# Mechanistic convergence of depression and suicidality on astrocyte fatty acid metabolism

**DOI:** 10.1101/2023.03.16.23287352

**Authors:** Eamon Fitzgerald, Nicholas O’Toole, Irina Pokhvisneva, Gustavo Turecki, Corina Nagy, Michael J Meaney

## Abstract

Genome-wide association studies (GWAS) show conceptual promise to identify novel mechanisms of major depressive disorder (MDD), but have not yet achieved this potential. One explanation is that MDD risk acts through complex expression networks, and GWAS-identified genes represent important components of these networks but in isolation are insufficient for their functional annotation. In this study, we aimed to identify and characterize the expression networks through which GWAS-identified MDD risk genes operate. We generated and characterized seeded co-expression networks of 252 MDD risk genes over 11 brain regions. We used principal component regression and Mendelian randomization to identify a relation between the networks of two such genes (*FADS1* and *ZKSCAN8*) and suicidal ideation. These networks were primarily expressed in astrocytes, enriched for functions related to fatty acid metabolism, and could define MDD-altered astrocyte states. We then identified *FGFR3* to *EPHA4* signaling as a putative downstream effector of these astrocyte states on synaptic function. Finally through transcriptomic and genetic analyses, we identify PPARA as a putative therapeutic target of these mechanisms in MDD. Our study defines a tractable pathway to translate genetic findings into therapeutically actionable mechanisms.

## Introduction

Major depressive disorder (MDD) is a leading contributor to the global burden of disease and is associated with immense socio-economic consequences^1,2^. Pharmacotherapy is typically the primary treatment strategy for MDD but large clinical trials show remission rates of up to 67% with common anti-depressant drugs^3^. Therefore, identifying novel therapies must be of the utmost priority, but this has been fundamentally hampered by our poor mechanistic understanding of MDD.

Genome-wide association studies (GWAS) show great conceptual promise to identify MDD mechanisms in an unbiased fashion. However, associating GWAS-identified loci to genes and downstream mechanisms has proven difficult. Transcriptome-wide association studies (TWAS) are a set of methods that attempt to annotate GWAS loci to the level of tissue-specific gene expression, by integrating GWAS summary statistics with expression quantitative trait loci (eQTLs). These methods have been widely applied and have implicated hundreds of genes in psychiatric disorders, such as MDD, but are hampered by false positives ^4,5^. TWAS method development promises to refine identified genes, but we suggest an integrative systems framework that leverages orthogonal methods and modalities is needed to confidently delineate causal pathways.

The genetic architecture of MDD is highly polygenic, and as such risk is not driven by individual genes, but through complex networks ^6,7^. Genetically identified risk genes are likely important components of these networks, but in isolation have a minimal contribution to overall risk ^7,8^. Co-expression networks are one method to identify the broad networks through which genes act. They are typically built in an unbiased fashion using RNA-sequencing datasets ^9^, however, an alternative method is to construct seeded co-expression networks, where networks are built around candidate genes. These seeded co-expression networks have been used successfully to interrogate the mechanistic underpinnings of psychiatric disorders ^10,11^. In this study, we hypothesized that generating and characterizing seeded co-expression networks for MDD TWAS genes would allow us to understand therapeutic relevant mechanisms of MDD.

We first systematically searched the literature to identify MDD TWAS genes. We then built 252 seeded co-expression networks, across 11 CNS regions using 1113 samples from the gene tissue expression (GTEx) study. We used principal component regression analyses followed by Mendelian randomization to identify a role for *FADS1* (*fatty acid desaturase 1*) and *ZKSCAN8* (*zinc finger with KRAB and SCAN domains 8*) co-expression networks in the prefrontal cortex with suicidal ideation. These networks functionally converged on fatty acid metabolism in astrocytes and network-based clustering of single nucleus RNA sequencing (snRNA-seq) data could define biased astrocyte states in MDD. We then identified *FGFR3* (*Fibroblast growth factor receptor 3*) to *EPHA4* (*EPH Receptor A4*) signaling as a putative downstream effector of these altered states on neuronal function. We finally identify PPARA (peroxisome proliferator activated receptor alpha) as a druggable regulator of these processes.

## Results

### Identification and characterization of MDD-seeded co-expression networks

We first systematically searched the literature to identify genes implicated in MDD by TWAS methodologies (see methods and **Figure 1**). This review highlighted 252 MDD TWAS genes that were present across regions in the GTEx v8 dataset (**Figure 2A**; **Supplementary table 1)**. As expected, the TWAS genes were strongly enriched in MDD GWAS summary statistics using MAGMA with different window sizes ^12^, but the strongest enrichment was achieved by H-MAGMA (P=3e-11) ^13^ (**Figure S1B**). We then asked if the TWAS genes were enriched within co-expression modules built using an unbiased approach (WGCNA; weighted correlation network analysis) from the CNS tissues of GTEx or a post-mortem suicide dataset^14,15^. We observed no enrichment among any of the modules (**Figure S1A**), suggesting a seeded approach may be better suited for mechanistic inference.

**Figure 1:**
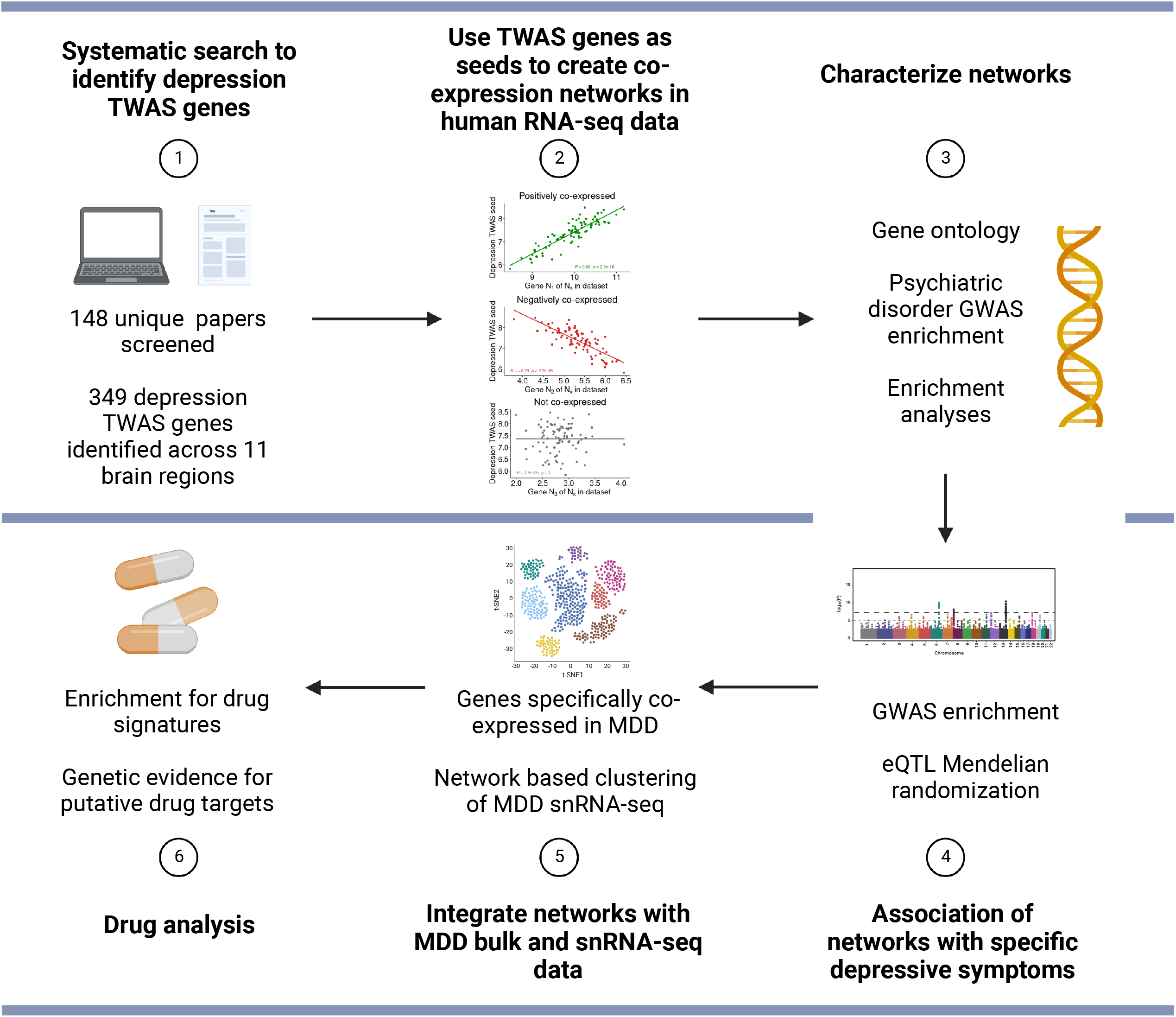
Schematic of the current study.

**Figure 2:**
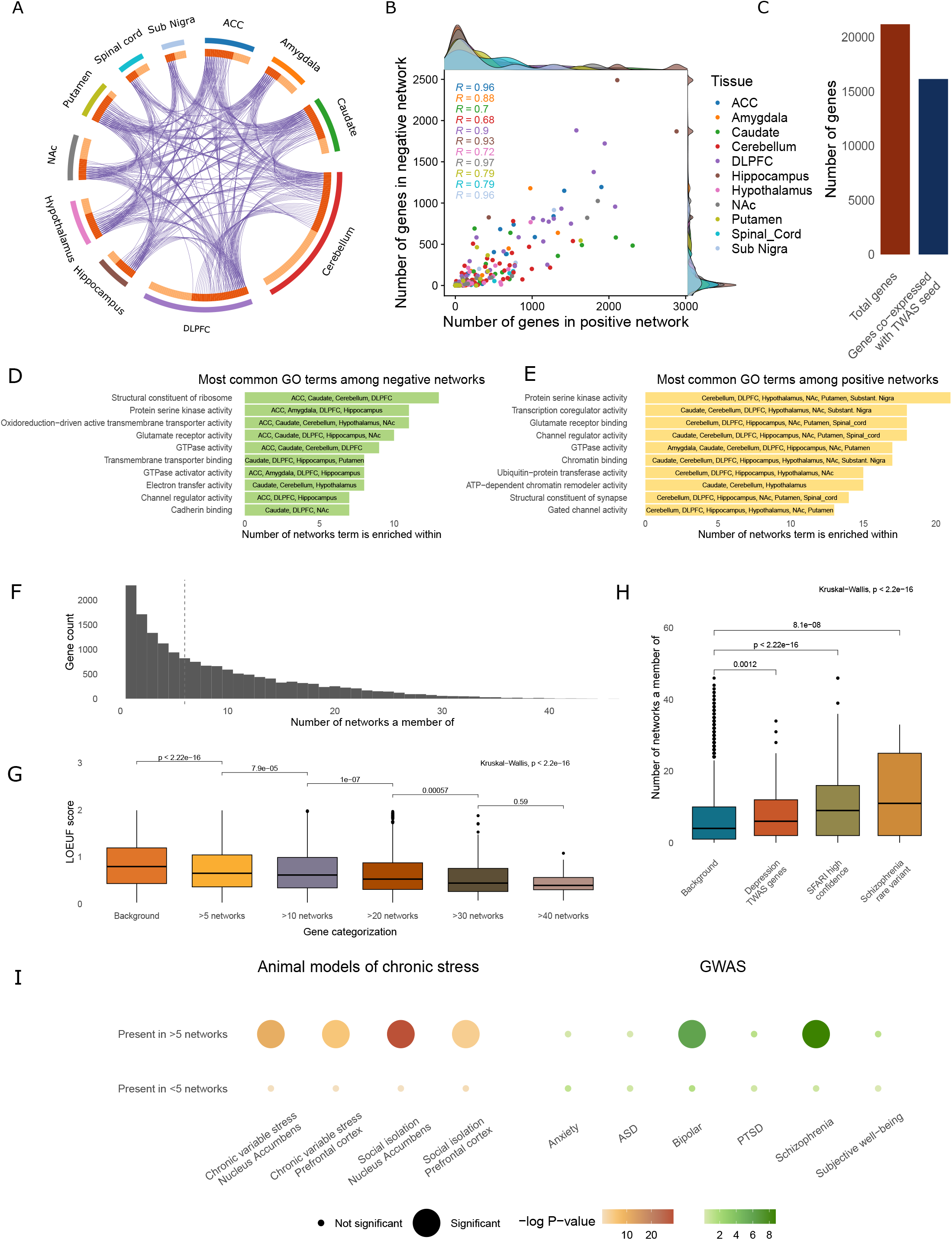
Generation and characterization of seeded co-expression networks. A. Circos plot describing the MDD TWAS seeds used to generate co-expression networks. Each outer segment represents an individual CNS region. The width of the segment is proportional to the number of seeds in that region. Light orange on the inner bar indicates a seed unique to that region and dark orange indicates that the seed is shared with another region, with this link represented by the purple connector. B. Scatter and density plots of all generated co-expression networks describing the number of genes positively and negatively co-expressed with each MDD TWAS gene. Pearson correlations between positive and negative networks for each region are shown. C. Bar plot representing the total number of genes in the full gene expression dataset (red bar) and those genes co-expressed with at least one MDD TWAS seed (dark blue bar). D and E. The most common GO terms enriched among the negative (D) and positive (E) networks. The x-axis is a count of the number of networks the relevant term is enriched within at an FDR P-value < 0.05. The regions with networks enriched for the relevant term are listed within each bar. F. Histogram of the number of networks genes co-expressed with at least one TWAS seed (also represented in the dark blue bar of C) are present within, includes both positive and negative networks. G. LOEUF scores for genes present in more than the indicated number of networks. Each category is inclusive of the subsequent categories i.e. genes present in >40 networks are also present in the >30 network category and so forth. Kruskal–Wallis test was first conducted (P<2.2e-16) followed by Wilcoxon post-hoc test (P-values indicated on graph). H. The number of networks MDD TWAS genes, SFARI high-confidence genes and genes associated with schizophrenia rare variants appear within. Kruskal–Wallis test was first conducted (P<2.2e-16) followed by Wilcoxon post-hoc test (P-values indicated on graph). I. Enrichment analysis of those genes appearing in > 5 or < 5 networks for genes differentially expressed in animal models of chronic stress (brown gradient; Fisher’s Exact Test) or in GWAS for psychiatric-related traits or disorders (green gradient; H-MAGMA). The larger dots indicate a corrected P-value < 0.05, with a darker colour indicating a lower P-value.

We then generated seeded co-expression networks for the 252 MDD TWAS genes in their relevant CNS tissue using the GTEx v8 dataset ^16^. Following filtering, normalization, and regression of technical covariates (see methods), we tested each MDD TWAS seed for co-expression with each of the 21194 genes in the dataset. Any gene with a correlation coefficient > 0.5 or < -0.5 and an FDR P-value < 0.05, was said to be positively or negatively co-expressed, respectively. Together all genes positively or negatively co-expressed with a particular seed were said to form positive or negative co-expression networks, respectively. There was a strong correlation across all regions between the size of the positive and negative co-expression networks (**Figure 2B**). Remarkably 76% of all genes tested for co-expression were co-expressed with at least one MDD TWAS seed (**Figure 2C**). Plasticity and activity-dependent processes were among the most common gene ontology (GO) terms enriched across all networks, with similar terms enriched in both positive and negative networks, suggesting oppositional regulation of processes by MDD risk genes (**Figure 2D and 2E**).

Of all co-expressed genes, 86% were co-expressed with more than one TWAS seed (**Figure 2F**) and there was a dose-dependent inverse relationship between the number of networks a particular gene was present within and their tolerance to mutation in the gnomAD database ^17^ (**Figure 2G)**. Psychiatric risk genes are typically highly intolerant to mutations, prompting us to ask if they were present in more networks than the background set of brain-expressed genes. Indeed we saw MDD TWAS genes, SFARI high-confidence autism genes, and schizophrenia risk genes (associated with rare variants ^18^) were present in more networks than expected (**Figure 2H**). We next categorized genes into those present in more or less than five networks for enrichment analyses. Genes present in more than five networks were enriched in both an MDD GWAS 19 (**Figure S1C**) and genes differentially expressed in models of chronic stress ^20^ (chronic variable stress-nucleus accumbens P=1.5e-11; chronic variable stress-prefrontal cortex P=2.5e-6; social isolation-nucleus accumbens P=1.1e-28; social isolation-prefrontal cortex P=1.9e-5; Fischer’s Exact Test) (**Figure 2I**). Genes present in more than five networks were also enriched in GWAS of common variants for bipolar disorder (P=7.5e-7) and schizophrenia (P=1.7e-9; **Figure 2I**).

### Association of large co-expression networks with specific MDD symptoms

MDD is a clinically heterogenous entity, which prompted us to ask if our co-expression networks are associated with specific MDD symptoms. To answer this question, we limited our search to networks of more than 750 genes (63 networks, top 12.5% in size), reasoning that larger networks would be more biologically relevant, as well as more amenable to functional characterization and downstream analyses. We used GWAS summary statistics from the patient health questionnaire nine (PHQ9) answered as part of the UK Biobank ^21^. The PHQ9 queries the current presence of the nine DSM depressive symptoms. We used H-MAGMA, to associate all 63 networks with each of the nine symptoms. We hierarchically clustered the scaled beta coefficients from the principal component regressions, which identified three groups of networks that were primarily associated with three groups of symptoms (**Figure 3A**).

**Figure 3:**
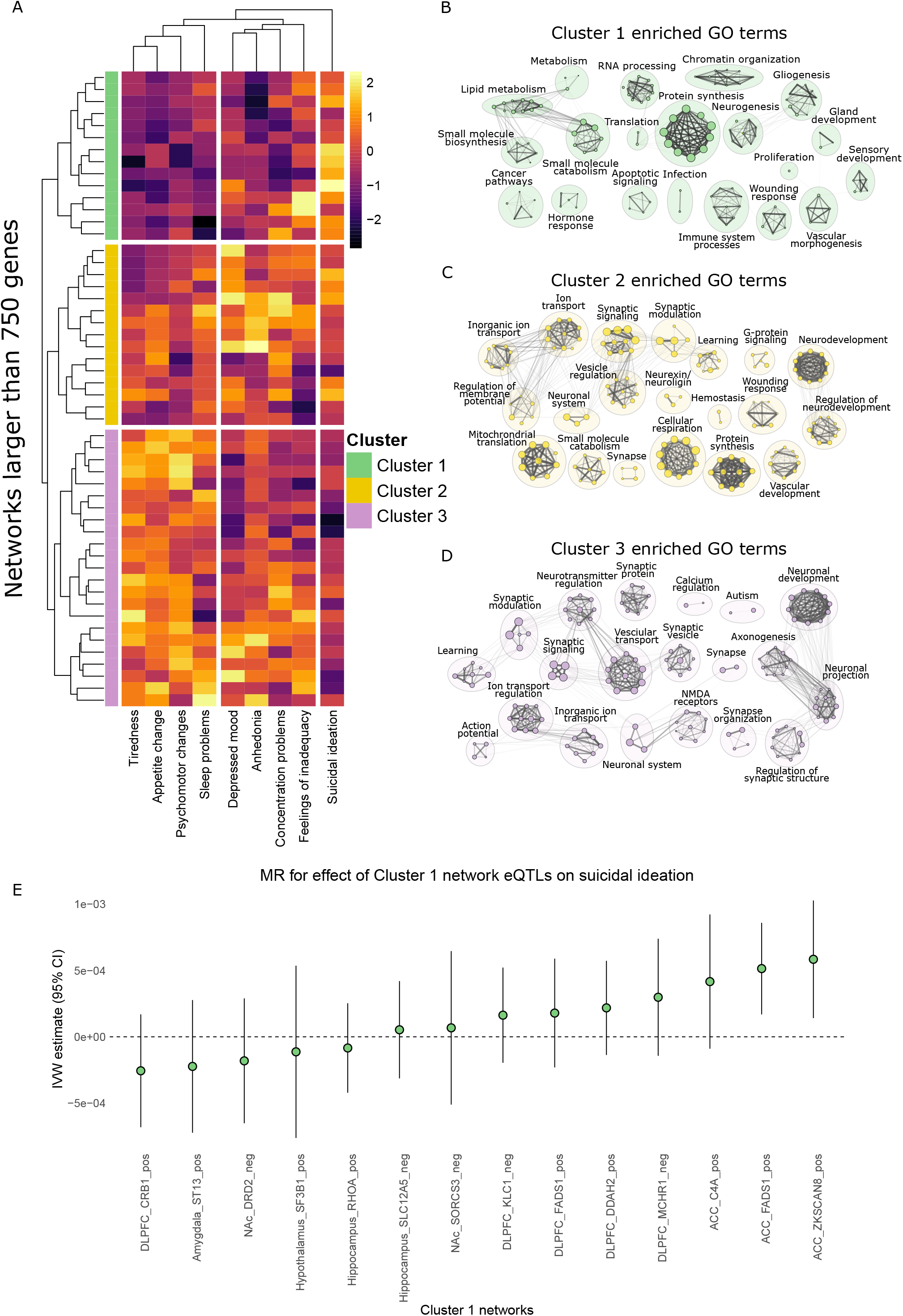
Clustering of co-expression networks by their enrichment in the nine DSM symptoms of MDD. A. Heatmap showing the enrichment of all networks (positive and negative) larger than 750 genes (y-axis) in GWAS summary statistics from PHQ9 ascertained symptoms of MDD (x-axis). The beta coefficient (row scaled) of the principal component regression for the effect of each network on each symptom is shown in the relevant tile. Yellow and black indicate a higher or lower beta coefficient, respectively. Results are grouped by hierarchical clustering into three symptom clusters and three network clusters. Dendrogram for network clusters is shown on the y-axis. B-D. All GO terms enriched (FDR corrected P-value < 0.05) for Cluster 1 (B; green colours), Cluster 2 (C; yellow colours) and Cluster 3 (D; purple colours) networks are grouped into nodes, with edges between nodes representing genes common between terms. Node size corresponds to Z-score of enrichment. Labels describe the broad annotation of their adjacent cluster of nodes. E. Mendelian randomization (MR) for an effect of eQTLs associated with Cluster 1 networks (x-axis) on suicidal ideation. Inverse variance weighted (IVW) effect size (y-axis) is shown with 95% confidence intervals.

Interestingly these three clusters are associated with functionally distinct GO enrichment patterns. Cluster 1 networks, whare ich primarily associated with suicidal ideation, were enriched for terms related to lipid metabolism, protein synthesis, inflammation and gliogenesis (**Figure 3B**). Cluster 2 networks, which primarily associated with dysphoric symptoms (anhedonia, depressed mood, concentration problems and feelings of inadequacy), was enriched for GO terms related to ion transport, synaptic processes, metabolism and neurodevelopment (**Figure 3C**). Cluster 3 networks, which primarily associated with somatic symptoms (tiredness, appetite changes, psychomotor changes and sleep problems), was enriched for GO terms related to neuronal plasticity and function (**Figure 3D**). Considering our poor understanding of suicidal traits and the tractability of studying them in post-mortem tissue we focused our subsequent analysis on the Cluster 1 networks

We used Mendelian randomization (MR) as an orthogonal method to associate Cluster 1 networks with suicidal ideation. We annotated the genes within each Cluster 1 network to independent eQTLs (from the GTEx catalogue ^16^), for use as instrumental variables. We then estimated the effect of these eQTLs on the suicidal ideation GWAS form the PHQ9. Using the inverse variance weighted method, two networks (FADS1 positive network in the anterior cingulate cortex and the ZKSCAN8 positive network in the anterior cingulate cortex, hereafter referred to as FADS1_ACC_ and ZKSCAN8_ACC_) had nominally significant P-values, with the FADS1_ACC_ network passing correction for multiple comparisons (FADS1_ACC_ P=0.003, FDR P=0.04, β=5.14e-04; ZKSCAN8_ACC_ P=0.009, FDR P=0.06, β=5.84e-04; Inverse variance weighted method; **Figure 3E**). These networks also had significant effects (P<0.05) with at least one other MR method and were directionally consistent across all methods (**Figure S2**), without evidence of instrument heterogeneity or horizontal pleiotropy (**Supplementary tables 2-4**). Interestingly *FADS1* was also a TWAS gene in the DLPFC, and this network (FADS1_DLPFC_) also clustered with suicidal ideation and was directionally consistent, although not significant (P=0.39), with a positive effect in the MR analysis. The DLPFC and ACC are also the brain regions with the strongest causal evidence for a relation with MDD ^22,23^ and targeting the interactions between the two regions is used therapeutically ^24^. Therefore, we chose to focus our subsequent analyses on the FADS1_ACC_, FADS1_DLPFC_ and ZKSCAN8_ACC_ networks (**Supplementary table 5**).

### The FADS1_ACC_, FADS1_DLPFC_ and ZKSCAN8_ACC_ networks converge on fatty acid metabolism in astrocytes

We first used a bootstrapping approach to estimate the size (i.e. number of co-expressed genes) of the three networks across all CNS regions present in the GTEx catalogue, which found the largest networks in the regions more commonly implicated in MDD (DLPFC, ACC, hippocampus and nucleus accumbens). Whereas regions less commonly associated with MDD, such as the cerebellum and spinal cord, had the smallest network sizes (**Figure S3A**).

We found a striking consistency across the three networks, with a strong congruence across correlation coefficients for all 21194 genes tested for co-expression (**Figure 4A**; R = 0.81 to 0.92; Pearson correlation) and within their co-expression networks (i.e. R > 0.5 and FDR P-value < 0.05; **Figure 4B**). All three networks were also enriched for similar GO terms, primarily relating to fatty acid metabolism (**Figure 4C**), and were also specifically expressed in astrocytes (**Figure 4D**). We also note that all three networks were strongly enriched for protein-protein interactions (**Figure S4**). *FADS1* is a fatty acid desaturase with a well characterized role in lipid metabolism ^25^. On the other hand, ZKSCAN8 is a transcription factor without an established role in fatty acid metabolism, but has been reported to bind to the *FADS1* locus ^26^, suggesting ZKSCAN8 may affect depression risk through *FADS1* regulation.

**Figure 4:**
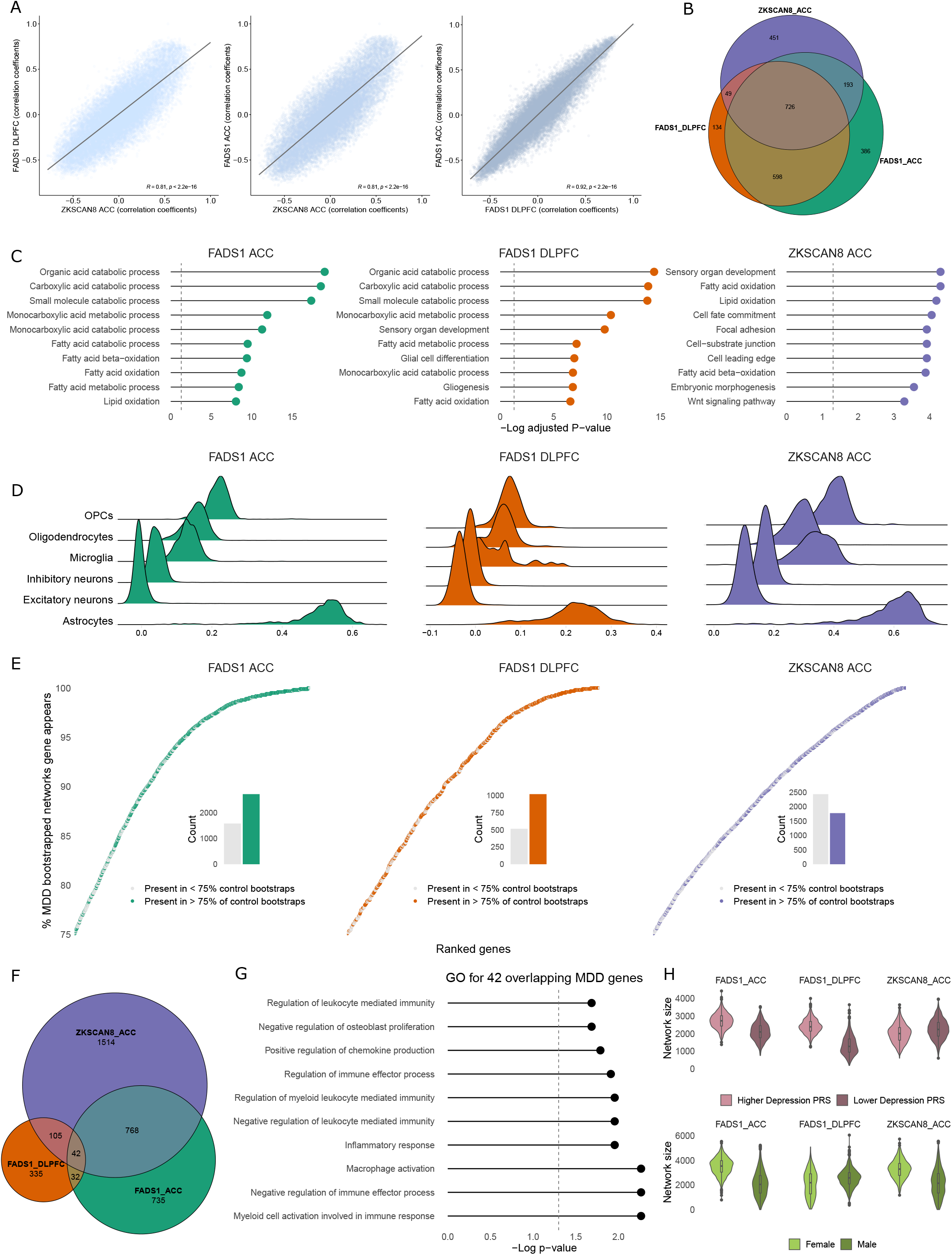
FADS1 and ZKSCAN8 networks in the prefrontal cortex converge on fatty acid metabolism in astrocytes. A. Scatter plots comparing the correlation coefficients of FADS1_ACC_, FADS1 DLFPC, and ZKSCAN8_ACC_ networks for all 21194 genes tested for co-expression. Pearson correlation coefficient and associated P-value is shown for each comparison. B. Venn diagram showing the overlap in genes within the FADS1_ACC_ (green), FADS1 DLFPC (orange) and ZKSCAN8_ACC_ (purple) positive networks. C and D. GO (C) and single cell expression (D) for the FADS1_ACC_ (green), FADS1 DLFPC (orange) and ZKSCAN8_ACC_ (purple) positive networks. E. MDD bootstrapped networks in the Labonté *et al* dataset for FADS1_ACC_ (green), FADS1 DLFPC (orange) and ZKSCAN8_ACC_ (purple). In each panel a dot represents a gene ranked by the percentage of 1000 bootstrapped networks it appears within. A grey colour indicates that the gene also appears in more than 75% of bootstrapped networks from control individuals. A green, orange or purple colour indicates that the gene is specifically co-expressed in MDD. Counts of these genes appear as insets. F. The overlap in MDD specific co-expressed genes across networks (i.e. the green, orange, or purple bars from the insets within E). G. GO terms enriched for the 42 common genes specifically co-expressed in MDD across the 3 networks. H. Bootstrapped network size in GTEx for the FADS1_ACC_, FADS1_DLPFC_ and ZKSCAN8_ACC_ based on MDD polygenic risk score (PRS; light and dark maroon; partitioned by median split) or sex (light green and dark green for females and males, respectively). 1000 bootstrap iterations were used for all size estimations.

We next asked if we could identify genes specifically co-expressed with FADS1_DLPFC_, ZKSCAN8_ACC_ and FADS1_ACC_ in post-mortem MDD tissue. We used a bootstrapping approach to construct networks from control and MDD individuals using bulk RNA-sequencing data from Labonté *et al* ^27^ (**Figure 4E**). This approach identified 335, 1514 and 735 genes in the FADS1_DLPFC_, ZKSCAN8_ACC_ and FADS1_ACC_ networks, respectively, which were specifically co-expressed in MDD. Considering the concordance of network function and cell-specificity among these three networks, we reasoned these networks were likely acting through a common mechanism. We accordingly identified 42 genes that were common across all three networks (**Figure 4F**), which were enriched for GO terms related to inflammation (**Figure 4G**). Mounting evidence suggests a core role for inflammation in MDD ^28^ and fatty acid signalling has important roles in inflammatory signalling ^29^, but the interplay with between fatty acid signalling and inflammation in MDD has not been well-studied.

Interestingly, in the GTEx dataset we also observed a notable increase in size of the FADS1_ACC_ and FADS1_DLPFC_ networks in individuals with a high MDD polygenic risk score (PRS) compared to a low MDD PRS, and an increased size of the ZKSCAN8_ACC_ and FADS1_ACC_ networks in females compared to males (**Figure 4H**), suggesting the three networks were also sensitive to depression risk factors. Of note, these changes occurred in the absence of *FADS1* or *ZKSCAN8* expression differences between either high/low MDD PRS or female/male groups (**Figure S3B**).

### FADS1_DLPFC_ network-based clustering identifies astrocyte states altered in MDD

Astrocytes have diverse functions, leading us to wonder if the networks could define astrocyte states and if these states would be altered in MDD. We answered this question using snRNA-seq data from the DLPFC of control and MDD individuals from Nagy *et al* ^30^. We clustered the astrocytes from this dataset based only on the genes present in the FADS1_DLPFC_ network (see methods section). This clustering approach allowed us to ask if variation in expression of the FADS1_DLPFC_ network could define specific astrocyte states and then ask if these states were altered in MDD. We identified three clusters of astrocytes using this approach (**Figure 5A and 5B**; **Figure S5A-S5C**). The highest expression of FADS1_DLPFC_ genes was in Cluster 0 and Cluster 1, with lower expression in Cluster 2 (**Figure 5C**). Our initial characterization showed large skews in the distribution of astrocytes in control and MDD samples (**Figure 5D**), which we also observed using a neighbourhood-based approach to differential abundance testing, implemented through the MiloR package that allowed us to regress the effects of age and batch from the analysis (**Figure 5E** and **Figure S5D**).

**Figure 5:**
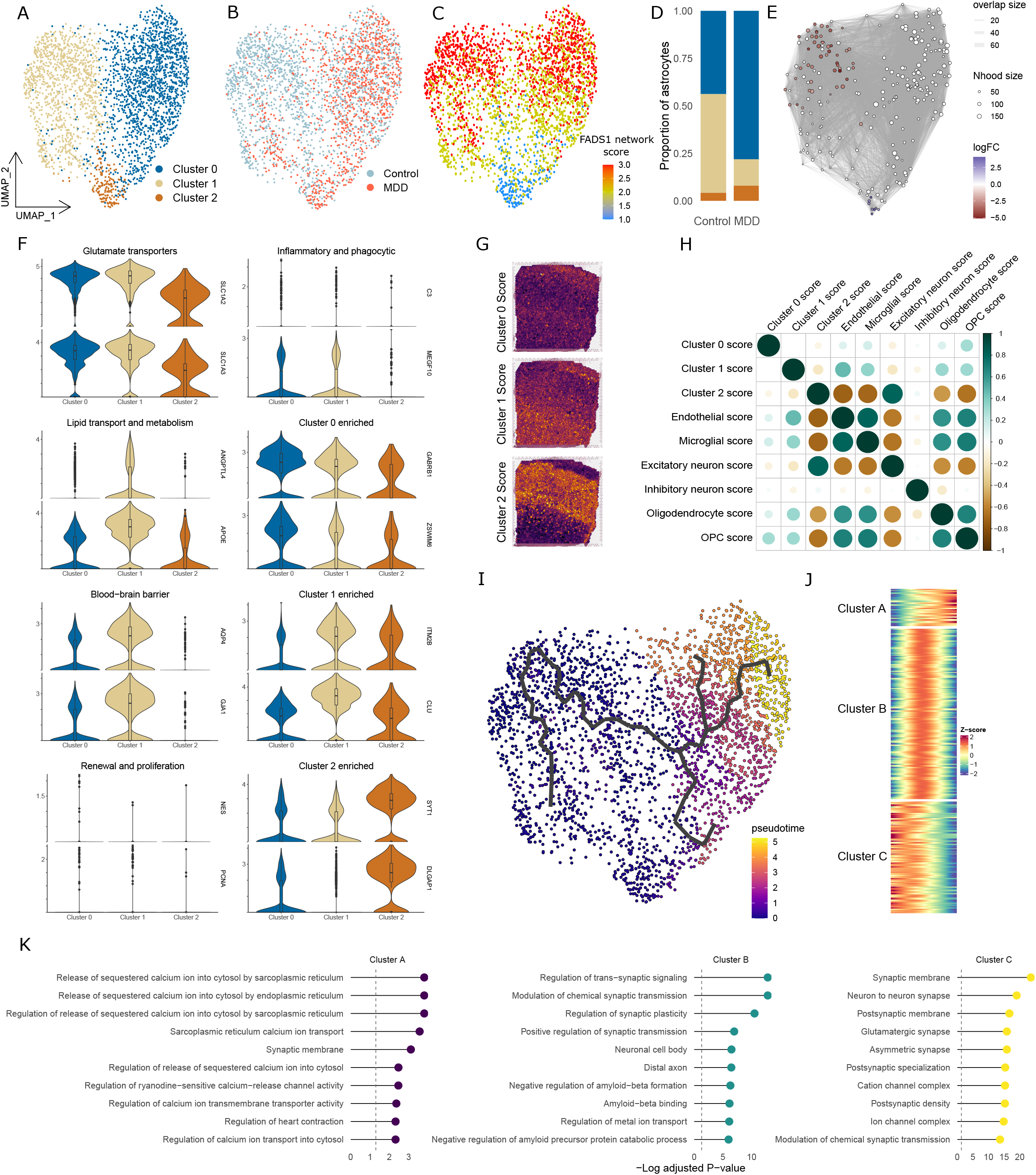
The FADS1_DLPFC_ network defines MDD biased astrocytic states. A-C. UMAP of FADS1_DLPFC_ network-based clustered astrocytes coloured by: defined clusters (A), control or MDD status (B) and a module score of the FADS1_DLPFC_ network (C; red and blue indicating higher or lower levels of the network, respectively). D. Stacked bar plot for the proportion of control and MDD nuclei in each cluster. E. Differential abundance of neighbourhoods in control and MDD groups adjusted for batch and age. Positive or negative logFC indicates an increased or decreased abundance in MDD, respectively. Edges between nodes represent nuclei common to neighbourhoods. F. Violin plots of gene expression across the three defined astrocyte clusters. G and H. Marker genes for the three astrocyte clusters projected into Visium data from the human DLPFC (Maynard *et al*). G displays representative images of the scores for the 3 clusters (layer I is at the top of the image, with the white matter at the bottom). The spatial correlations of these scores with scores generated in a similar manner for glia and neuronal populations is shown in J (average from 12 DLPFC slices). I. Pseudotime trajectory is plotted on the UMAP coordinates. J. The top 300 genes differentially expressed across pseudotime identifies 3 clusters (Cluster A, Cluster B and Cluster C). K. Gene ontology enrichment of the 3 clusters from J.

All three clusters expressed canonical astrocyte markers (**Figure 5F**). Cluster 1, which was sharply decreased in MDD, had strong expression of blood-brain barrier genes (BBB) (**Figure 5F**) and GO terms related to protein translation (**Figure S5G**; **Supplementary table 6**).

Cluster 2, which was increased in MDD, expressed a high level of synaptic genes (**Figure 5F**) and markers of this cluster were enriched for proteins present at the tripartite synapse (**Figure S5E**), in a GWAS for MDD (**Figure S5F**) and for GO terms related to synaptic function (**Figure S5G**; **Supplementary table 6**).

Cluster 0 seemed to fall between these clusters, expressing BBB and synaptic genes at a moderate level. We found GABRB1 (*Gamma-Aminobutyric Acid Type A Receptor Subunit Beta 1*), along with several ion channels (**Figure 5F**), among the genes most highly enriched in Cluster 0 suggesting a sensitivity to neuronal activity. Interestingly the GABA-A receptor, of which the *GABRB1* encodes a subunit, is the target for the newly approved anti-depressant brexnanolone ^31^. Furthermore, marker genes from Cluster 0 were also enriched in astrocytic components of the tripartite synapse (**Figure S5E)**, in a GWAS for MDD (**Figure S5F**) and GO terms related to cell-cell interactions (**Figure S5G**; **Supplementary table 6**).

We next wanted to assess the spatial localization of these astrocyte states and thereby uutilizedthe human DLPFC spatial transcriptomic dataset from Maynard *et al* ^32^. We used genes enriched (positive logFC and FDR P-value<0.05) within each astrocyte cluster, as well as genes enriched in broad classes of other canonical cell types, to build module scores in the spatial transcriptomic data (representative scores of the astrocyte clusters can be seen in **Figure 5G**) of 12 sections and correlated the spatial localisations of these scores. In line with our previous analysis, there was a strong colocalization between the Cluster 2 score and a score from excitatory neurons. Cluster 1 was also strongly expressed in layer I and the white matter, and was strongly colocalized with the endothelial cell score, again suggesting a role at the blood-brain barrier. Cluster 1 was also enriched for an astrocyte layer I signature previously observed in mice ^33^ (**Figure S5E**). The Cluster 0 score showed a diffuse pattern without strong colocalization for other cell types. Again suggesting it represented a functionally diffuse astrocyte state.

There was a sharp decrease in Cluster 1 nuclei in MDD, leading us to wonder if we could identify a transitional trajectory away from this cluster. To do this we used a pseudotime analysis employed within the Monocle 3 package, which identified a trajectory from Cluster 1 toward Cluster 0 (**Figure 5I**). We conducted differential expression along this trajectory and used K-means clustering to sort the top 300 differentially expressed genes into three clusters (**Figure 5J**; **Supplementary table 7**). Cluster A contained genes that were increased in their expression towards the end of the trajectory, with Cluster B and Cluster C genes primarily expressed in the middle and at the start of the trajectory, respectively. All three clusters were enriched for GO terms related to synaptic function (**Figure 5K**), suggesting astrocyte-neuronal communication may be an important regulator of astrocyte state transitions in MDD.

### FGFR3-EPHA4 as a driver of altered astrocyte-neuronal communication in MDD

We next wondered what lay downstream of these altered astrocyte states. We reasoned that the most likely metric of the downstream processes associated with genes *positively* correlated with FADS1 in the DLPFC was genes *negatively* correlated with *FADS1* in the DLPFC (hereafter the FADS1_DLPFC_ negative network). The FADS1_DLPFC_ negative network was primarily expressed in excitatory neurons (**Figure 6A**) and enriched for GO terms relating to the presynapse (**Figure 6B**). We also note there was a high degree of overlap between the FADS1_DLPFC_, FADS1_ACC,_ and ZKSCAN8_ACC_ negative networks (**Figure S6A**). The FADS1_DLPFC_ negative network was broadly expressed across all excitatory neuron subclusters, implying it was representative of a general excitatory presynaptic expression program (**Figure 6C**). Clustering of excitatory neurons based on genes within the FADS1_DLPFC_ negative network produced similar results to standard clustering (**Figure S6B-6E**).

**Figure 6:**
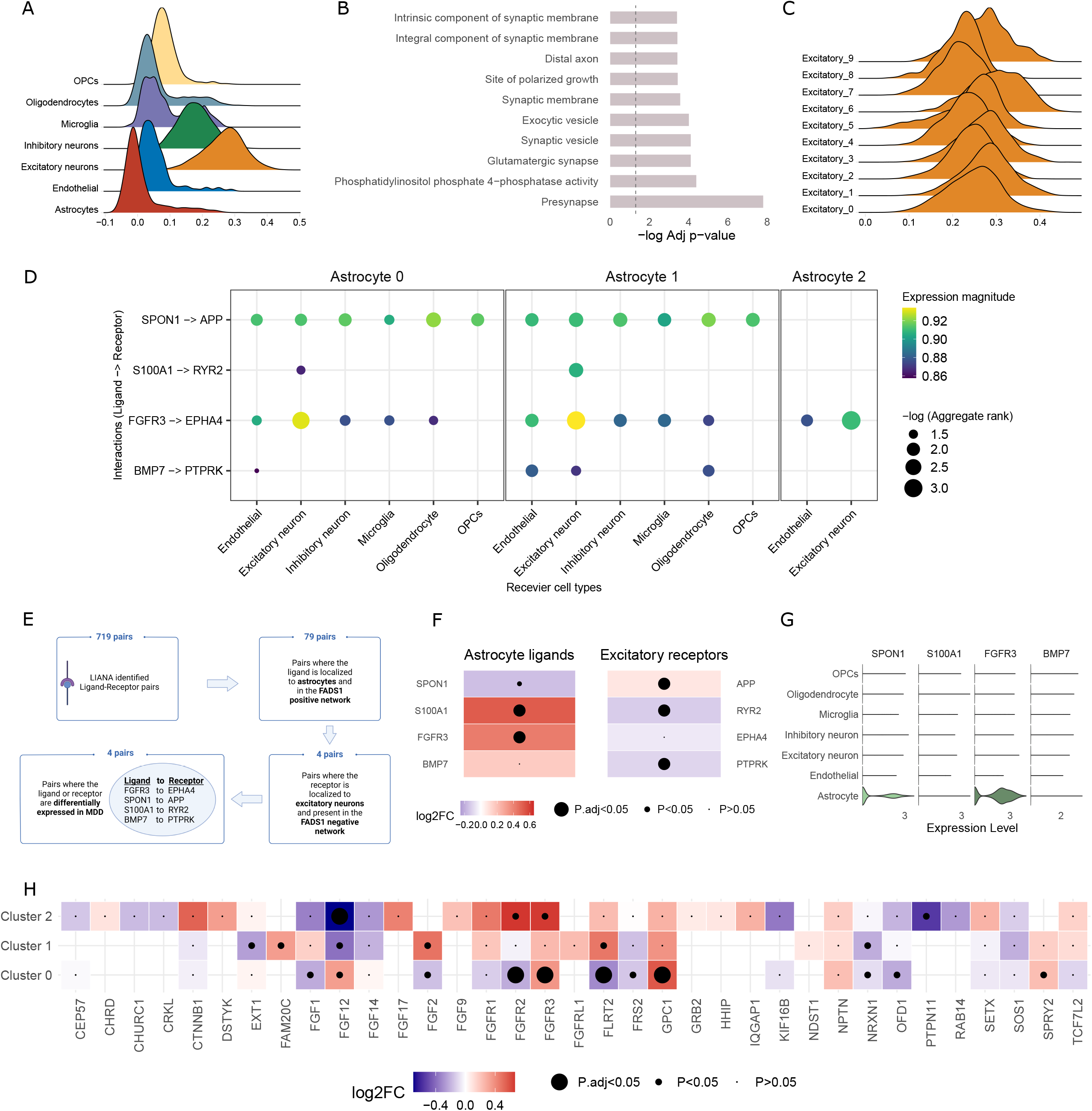
FGFR3 to EPHA4 signalling as a mediator of astrocyte-neuronal communication in MDD. A. The cell type specificity of the FADS1_DLPFC_ negative network module score. B. GO terms enriched in the FADS1_DLPFC_ negative network. C. Expression of the FADS1_DLPFC_ negative network in excitatory neuron clusters. D. Cell-cell interaction analysis using LIANA. Astrocytic expressed ligands are on the y-axis, with receptors on the x-axis. Colour of the dots represent the magnitude of expression, yellow representing higher expression of both ligand and receptor in their respective cell types. The size of the dots is proportional to the -log of the aggregate rank, which has an equivalent interpretation to a -log P-value. E. Schematic for identifying ligand-receptor pairs of interest. F. Heatmap showing the differential expression of the four LIANA nominated ligands in all astrocytes and the expression of their four putative receptors in all excitatory neurons. Tiles are coloured by the logFC of a comparison between control and MDD nuclei, with red indicating increased expression in MDD. The largest dot indicates an adjusted P-value < 0.05, the intermediate sized dot indicates an unadjusted P-value < 0.05 and the smallest dot indicates an unadjusted P-value > 0.05. P-values are based on Wilcoxon rank sum test comparison of gene expression comparing all control or MDD astrocytes or excitatory neurons. G. Cell type expression of the four astrocyte ligands prioritized by the LIANA analysis. H. Heatmap for differential expression of FGF receptor signalling pathway members (GO:0008543) in astrocyte clusters. Tiles are coloured by logFC, with red indicating increased expression in MDD and a larger dot indicating and adjusted P-value < 0.05. P-values are based on Wilcoxon rank sum test comparing expression between control and MDD nuclei within the specific astrocyte cluster.

We next employed LIANA, a consensus cell-cell interaction method ^34^, to identify complexes involved in communication between cell types. For this analysis, we used our three astrocyte states as the sending cell types and designated the broad cell classifications as the receiving cell types (**Figure 6D and 6E**). We then subset the ligand results to genes present in the FADS1_DLPFC_ *positive* network and the receptors to genes present in the FADS1_DLPFC_ *negative* network (**Figure 6D**). Considering the negative network was primarily expressed in excitatory neurons, we focused our attention on these nuclei and further refined our analysis to those ligand-receptor pairs with at least one member differentially expressed in MDD astrocytes or excitatory neurons, respectively (**Figure 6F**). This strategy produced a list of four ligand-receptor complexes; *SPON1* (*Spondin 1*) to *APP* (*Amyloid-beta precursor protein*), *S100A1* (*S100 Calcium Binding Protein A1*) to *RYR2* (*Ryanodine receptor 2*), *FGFR3* (*Fibroblast growth factor receptor 3*) to *EPHA4* (*EPH Receptor A4*) and *BMP7* (*Bone morphogenetic protein 7*) to *PTPRK* (*protein tyrosine phosphatase receptor type K*). We next examined the cell type expression of the astrocyte ligands and observed only minimal expression of *S100A1* and *BMP7* in astrocytes, with moderate expression of *SPON1* that was specific to astrocytes and high levels of *FGFR3* expression that was highly selective for astrocytes (**Figure 6G)**. *FGFR3* is a well-conserved regulator of astrocyte morphology ^35–37^, with several previous studies also reporting differential expression of *FGFR3* in post-mortem MDD tissue ^38–40^. Furthermore, EPHA4 is present at the synapse in the mouse brain ^41,42^, increases at the protein level in post-mortem MDD tissue ^43^, knockdown or inhibition shows anti-depressant-like effects in mice ^43,44^, has an established role in astrocyte-neuronal communication ^42^ and binds to FGFR3 ^45^. Therefore, *FGFR3-EPHA4* represented the best candidate to mediate altered astrocyte-neuronal communication in MDD. Several previous studies have broadly implicated FGF signaling in MDD^46,47^, prompting us to assess differential expression across the full FGF signaling family. Notably, we observed several FGF members were differentially expressed in astrocytes, suggesting altered *FGFR3-EPHA4* signaling may exist within the context of broad dysregulation of FGF signaling in MDD.

### PPARA as a therapeutic target in MDD

We next sought to identify druggable therapeutic targets from our analyses (**Figure 7A**). We reasoned two of our approaches were the most promising signatures to investigate; the 42 common genes that were specifically co-expressed in MDD (**Figure 4E and 4F**) and Cluster A genes that were upregulated towards the end of our pseudotime trajectory (**Figure 5J**). We analyzed these signatures for an association with the DrugMatrix and Connectivity Map databases of transcriptomic perturbations ^48^. We found 152 and 52 compounds associated with the MDD network specific and pseudotime lists, respectively (**Figure 7B** and **Supplementary table 8**). Considering the distinct nature of our signatures, we found a surprisingly strong convergence between results, with the PPARA agonist fenofibrate among the top hits for both signatures. We note these analyses define agents that modulate our signatures and may therefore be pro- or anti-depressant. For instance among the top hits was the pro-inflammatory agent, lipopolysaccharide (LPS), whose pro-depressive effects are well characterized ^28^. We also identified several interesting drug classes including anti-inflammatory (e.g. ampiroxicam and dexamethasone), sex-hormone targeting (e.g. diethylstilbestrol, norethindrone, tamoxifen and epitiostanol) and a selective-serotonin reuptake inhibitor (sertraline).

**Figure 7:**
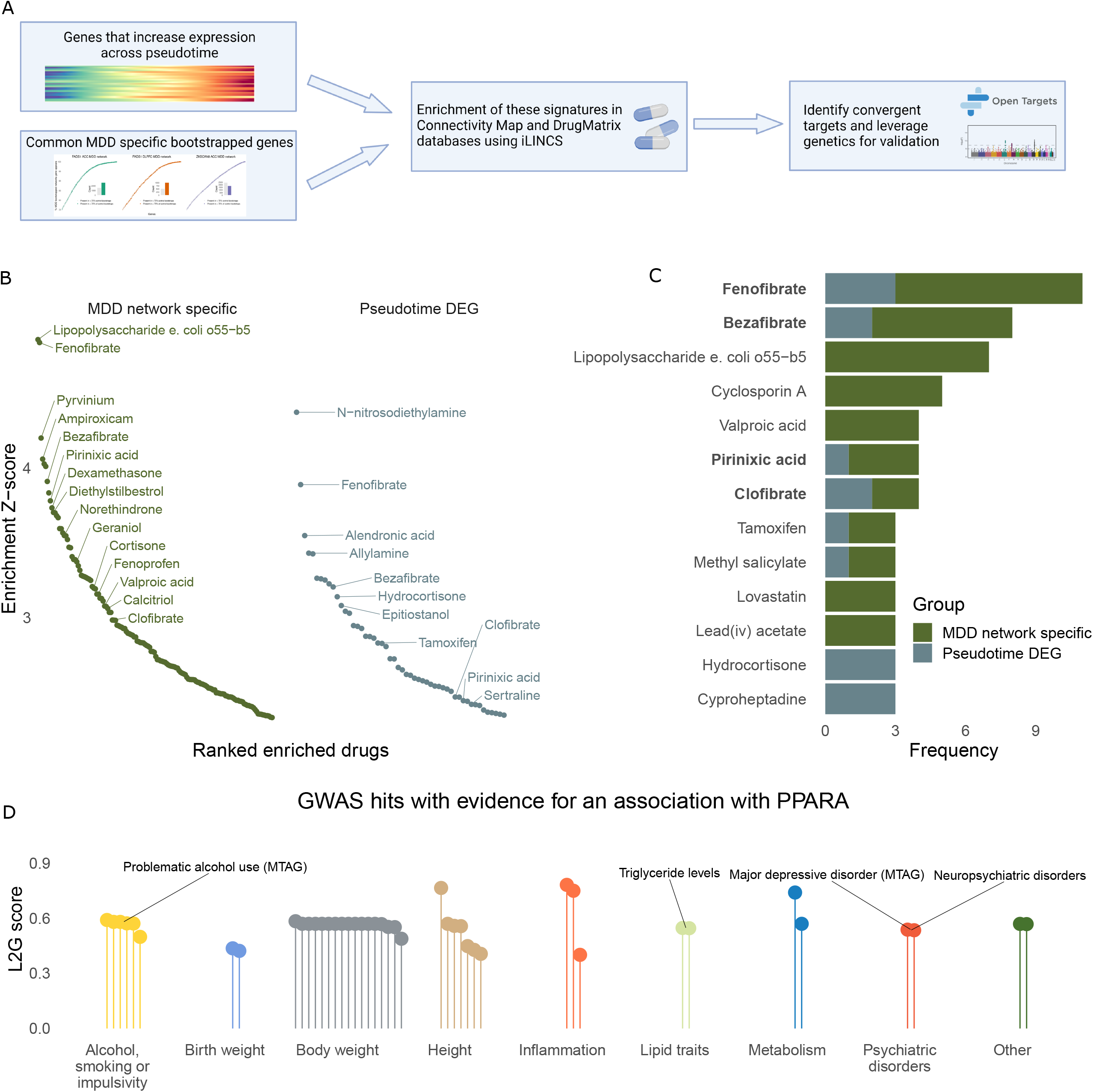
PPARA is a putative therapeutic target in MDD. A. Schematic of drug target prioritisation strategy. B. Significantly associated perturbagens from the DrugMatrix and Connectivity Map databases ranked by absolute Z-score for an association with common MDD network specific genes (dark green) or Cluster A pseudotime differentially expressed genes (dark blue). Notable perturbagens are labelled. Z-scores are generated from two-tailed P-values for the change in expression of the relevant signatures in the relevant perturbagen gene expression dataset. C. Count of perturbagen appearances (e.g. from different cell types or concentrations) from B. Drugs targeting PPARA are highlighted in bold. D. GWAS hits with evidence of an association with *PPARA* in the open targets database. The y-axis indicates the L2G (locus to gene) score, which ranges from 0-1 and can be interpreted as probability of being the causal gene under identical conditions to the GWAS training set. Each ‘lollipop’ on the x-axis represents a distinct GWAS, which are functionally grouped and notable GWAS are labelled.

These databases contain signatures across multiple cell lines and concentrations, therefore, we reasoned that an association across multiple conditions would provide stronger evidence of an association. Among the most frequently associated drugs were four that target PPARA. Moreover, each of the four drugs were associated with both the MDD network-specific and pseudotime DEG signatures (**Figure 7C**). PPARA is a nuclear receptor that regulates the expression of a range of lipid-modulating genes ^49^ and so represented an intriguing candidate. Encouragingly PPARA was also a member of the FADS1_ACC_ (R=0.79), FADS1_DLPFC_ (R=0.74)and ZKSCAN8_ACC_ (R=0.65) networks (**Figure S7A**), but the other PPAR family members *PPARD* and *PPARG* were not present in any of the three networks (**Figure S7B and S7C**).

We finally used the Open Targets platform to search for genetic associations with PPARA. Surprisingly there was a GWAS significant locus 88Kb from *PPARA* in a GWAS of MDD (**Figure 7D**; **Supplementary table 9**) ^50^, making it the second closest gene to the locus (WNT7B was the closest at 84Kb). We could not identify any brain QTLs for PPARA, despite its robust expression, which negated the possibility of confirming the association using colocalization or Mendelian randomization. There was however also evidence for *PPARA* as the causal gene in multiple other GWAS, including several pertaining to co-morbidities or risk factors for MDD including addiction, inflammation and body weight (**Figure 7D**). Together these data support the further study of PPARA as a putative therapeutic target in MDD.

## Discussion

In this study, we used multiple complementary and orthogonal methods to derive therapeutically actionable MDD mechanisms from genetic studies. We identify multiple expression networks associated with MDD and suicidality that converge on fatty acid metabolism within astrocytes of the prefrontal cortex. Furthermore, we identify PPARA as a putative anti-depressant target to modulate these mechanisms. Considering the diffuse definitions of MDD used in GWAS ^51^ and the limitations of TWAS ^5^, we illustrate that integrating results across data modalities and using orthogonal methods is a viable strategy for the delineation of therapeutically actionable mechanisms in MDD.

Several streams of research have linked astrocyte alterations to MDD in humans ^52–54^ or depressive-like behaviors in animal models ^55–57^. One notable finding from these studies has been a specific disruption of astrocyte function at the BBB ^55,56,58,59^. We could observe similar effects by clustering astrocytes based on the FADS1_DLPFC_ network, suggesting a role for fatty acid metabolism in astrocyte dysregulation at the BBB. Many fatty acids, particularly polyunsaturated fatty acids (PUFAs), are supplied to the brain via the periphery ^29^. PUFAs are metabolized by FADS1 into various active metabolites, such as arachidonic acid and eicosapentaenoic acid, which play an important role in the regulation of inflammation ^60^. Interestingly we found genes specifically co-expressed with the three networks in MDD were enriched for inflammatory processes, suggesting PUFA metabolism is a potential contributing mechanism to altered inflammatory signaling in MDD. Furthermore, both FGFR3 and EPHA4, to whom we ascribe a putative role for mediating downstream synaptic effects of the FADS1_DLPFC_ network, have characterized roles in inflammatory signaling pathways ^61–63^. Therefore, our study suggests a model whereby astrocyte dysfunction at the BBB facilitates the entry of PUFAs to the brain, which are metabolized by FADS1 to produce inflammatory mediators that lead to altered astrocyte-neuronal communication. Crucially PUFAs also act as endogenous agonists for PPARA ^29^, suggesting modulation of this process may be therapeutically relevant.

Our study points to PPARA as a putative therapeutic target in MDD, with the literature suggesting PPARA agonism as the likely desired therapeutic action. For instance, administration of palmitoylethanolamide (PEA; an endogenous PPARA ligand), or the inhibition of PEA degradation, produces anti-depressant-like effects in mice ^64–66^ and PEA improved the anti-depressant effect of Citalopram when used as an adjuvant in a small clinical trial ^67^. Non-endogenous PPARA agonists such as fenofibrate, WY14643 and aspirin also have a range of PPARA-dependent effects within the rodent brain including anti-depressant-like ^65,68,69^ and anxiolytic-like effects ^65,70^, modulation of memory ^70–72^, altered astrocyte function ^70,73^ and increased synaptic plasticity ^71,72^. Furthermore, knocking out PPARA in mice results in reduced cognitive flexibility and altered synaptic gene expression ^74–76^. Together with these data, our results provide a strong basis for the further investigation of PPARA agonists as anti-depressant agents.

Currently, the primary clinical use of the PPARA agonists is for dyslipidemia ^77^ and interestingly peripheral lipid alterations are commonly observed in MDD ^78–80^. The *FADS* locus (includes *FADS1, FADS2* and *FADS3*) is the most pleiotropic regulator of peripheral metabolite levels, with significant genetic associations for 79 metabolites, including 75 lipid species ^25^. Furthermore, there is also evidence to suggest that peripheral lipid dysregulation caused by the FADS locus is causal in MDD ^81^. FADS1 has also been identified through TWAS as a depression risk gene within the brain and we describe alterations in FADS1 co-expression networks in the prefrontal cortex associated with MDD and suicidality. Disentangling the peripheral and central effects of FADS1 lipid metabolism in MDD is an important future question. To fully address this question, larger cohorts and emerging methods with improved throughput and resolution for lipid analysis that are suitable for use in post-mortem tissue will be essential.

The current study also comes with limitations. First, the generation of robust co-expression networks requires large sample sizes and study designs, such as ours, will benefit from increasing MDD cohort sizes. Secondly, we also rely on transcriptomic methods throughout this study. However, these methods are limited in their ability to infer events that may not be well represented at the transcriptome level, such as lipid metabolism. Emerging single-cell metabolomic methods ^82^ promise to build upon our results.

In conclusion, we implicate fatty acid metabolism in astrocytes as a driving mechanism in MDD and suicidality and identify PPARA as a putative therapeutic target to modulate this pathway.

## Methods

### Systematic search

We conducted a systematic search of the literature to identify previous studies that used a TWAS framework to identify MDD risk genes. On March 14^th^, 2022 we searched the PubMed and Web of Science databases for manuscripts containing [“MDD” or “Major depressive disorder”] AND [“Transcriptome-wide association study” or “TWAS”] from the previous five years. Only TWAS were considered, and proteome-wide association studies were removed. Duplicate studies were removed by matching PMIDs and the titles of 148 unique articles were then screened for relevance, returning 87 articles whose abstracts were evaluated for relevance. After this process 23 studies remained for full text evaluation, of which 7 were deemed suitable. See **Supplementary Table 1** for a list of included studies. TWAS genes carried forward were limited to GTEx CNS tissues to fit within the scope of the analyses, leaving 359 MDD TWAS genes across 11 CNS regions. TWAS genes identified in the GTEx “Cerebellum” or “Cerebellar hemispheres” were combined as were those in “Frontal cortex BA9” and “Cortex”, as the GTEx consortium notes these as technical replicates.

### Bulk RNA sequencing quality control and normalisation

#### Gene Tissue Expression (GTEx) consortium

GTEx v8 data ^16^ were downloaded from the GTEx portal or through dbGaP (accession number phs000424.v9.p2). Raw data were filtered to retain genes expressed in all brain regions, to facilitate regional comparisons. Only genes and samples passing GTEx quality control were used in the downstream analysis. The cortical and cerebellar samples processed through alternative methods to the rest of the samples were removed. Samples were excluded if they were ineligible for the study or the presence of the following was noted: Amyotrophic Lateral Sclerosis, Alzheimer’s OR Dementia, Dementia With Unknown Cause, Active Encephalitis, Influenza (acute viral infection including avian influenza), Multiple Sclerosis, Parkinson’s Disease, Reyes Syndrome, Documented Sepsis, Systemic Lupus, Cerebrovascular Disease (stroke, TIA, embolism, aneurysm, other circulatory disorder affecting the brain), Alzheimer’s, Bacterial Infections (including septicaemia, meningococcal disease, staphylococcal infection, streptococcus, sepsis), Current Diagnosis Of Cancer, MDD, Heroin Use, Prescription pill abuse, Pneumonia, Schizophrenia, Unexplained seizures or Positive blood cultures. Data were normalized using DESeq2 v1.34.0 with the TMM method and log normalization. Genes were filtered to those with > 0.1 CPM in >25% of samples. Outlying samples were identified by a connectivity Z-score greater than three standard deviations and were removed. There was no effect of RIN on mapped reads (linear regression P-value = 0.979). A principal component analysis of 32 sequencing metrics was conducted and the first 10 principal components along with RIN, interval of onset to death, Hardy scale, body refrigeration, sex and age were regressed from the data using a linear mixed effects model with subject as a random intercept term with the lme4 v1.1-31 package ^83^.

#### Labonté et al

Data from Labonté *et al* ^27^ were downloaded from GSE102556 in FPKM form. Genes were filtered to those with an FPKM > 0.5 in 25% of samples, leaving 22893 genes in the dataset. The effects of age, age^2^, pH, sex, post-mortem interval, RIN and RIN^2^ were then regressed from the data.

### Generation of co-expression networks

Co-expression networks were generated using a Pearson correlation between the TWAS gene and all genes expressed in the dataset. A gene was said to be positively co-expressed with an R > 0.5 and an FDR P-value < 0.05, a gene was said to be negatively co-expressed with an R < -0.5 and an FDR < 0.05. Bootstrapping was carried out using 1000 iterations while down sampling to the smallest sample size among compared groups.

### Mutational intolerance analysis

Loss of function metrics v2.1.1 were downloaded from the gnomAD database ^17^. We used the LOEUF score as a conservative measure of mutational intolerance. All genes expressed in the GTEx dataset that were tested for co-expression were used as background in the analysis.

### Enrichment analyses

Fisher’s Exact Test was conducted to test enrichment between gene sets of interest and co-expression networks as implemented in the GeneOverlap v1.30.0 package ^84^. All genes expressed in the GTEx dataset that were tested for co-expression were used as background in the analysis.

### GWAS enrichment

We conducted GWAS enrichment using MAGMA v1.10 ^12^, with GWAS annotated to the gene level either using H-MAGMA ^13^ or the standard window approach in MAGMA. We compared the H-MAGMA approach to the window approach using enrichment of MDD TWAS genes in a MDD GWAS. H-MAGMA returned stronger statistical results and was therefore used for the remainder of the analysis. H-MAGMA is an extension to the MAGMA framework under which the GWAS of interest is annotated to the gene level using Hi-C data from a relevant region. We used SNP annotations from the 1000 genomes European dataset and gene annotations from the NCBI website build 38. We annotated GWAS using Hi-C data from the DLPFC ^85^, which was obtained from https://zenodo.org/record/6382668#.Y9PSG3bMJPY.

For the PHQ9 analysis, the beta coefficients from the principal component regression were scaled for each network and hierarchically clustered using the pheatmap v1.0.12 package ^86^. The dendrograms were then cut to form three clusters of rows and three clusters of symptoms.

For details of GWAS summary statistics used and their sources see **Supplementary Table 10**.

### Gene ontology (GO) analysis

We conducted GO analysis using the clusterProfiler v4.2.2 package ^87^, with all genes expressed in the relevant dataset used as background in the analysis. We ran enrichment for biological processes, cellular components, and molecular functions together. For Figure 3B-2D, significant terms were imported into Cytoscape v3.8.0 ^88^, where similar terms were clustered and a word cloud was used to suggest words common to all terms within a given cluster. We then manually refined these terms for brevity and clarity.

### Mendelian randomization

We conducted Mendelian randomization using eQTLs as instrumental variables for an effect on suicidal ideation. We obtained independent eQTLs from the GTEx v8 catalogue for all CNS relevant brain regions ^16^. We converted SNP location to rsIDs using the lookup tables provided by GTEx. We used the TwoSampleMR v0.5.6 package ^89^ to harmonize the effect alleles and analyse the effect of the eQTLs on suicidal ideation using the Inverse variance weighted, weighted median, weighted mode and MR Egger methods. We used leave-one-out and Cochranes Q-test to assess heterogeneity among SNPs and the Egger intercept to asses horizontal pleiotropy, all using standard settings in the TwoSampleMR package.

### Protein-protein interactions

Estimates of protein-protein interactions were made using the STRING database ^90^ using standard settings. Protein-protein interaction networks were then exported to Cytoscape v3.8.0 ^88^ for plotting.

### Single cell expression of networks

We assessed the single cell expression of co-expression networks using the module score function in Seurat v4.3.0 ^91^. We first downloaded single cell or single-nucleus data from the ACC and DLPFC (Tran *et al* ^92^ and Nagy *et al* ^30^, respectively). We downloaded the normalized ACC dataset as detailed in the Tran *et al* manuscript, and converted the Single Cell object to a Seurat object using the as.Seurat() function and identified variable features and principal components using the standard Seurat workflow. We retained the original clusters from this dataset, but combined them to form broad cell-type classes (e.g. astrocytes, oligodendrocytes, excitatory neurons, etc). We then generated a module score for plotting using the AddModuleScore() function in Seurat.

We downloaded the Nagy *et al* raw data from GSE144136 and used the cell type assignations from the original study, but again grouped them to from broad cell classes. We excluded nuclei expressing fewer than 700 and greater than 7000 genes, before normalization with the NormalizeData() function. Data were then split and integrated by batch using the Seurat method of data integration and the standard Seurat pipeline before using the AddModuleScore() function to identify cell type specific expression patterns.

### Analysis of snRNA-seq data

To cluster astrocytes based on variation in expression of genes within the FADS1_DLPFC_ networks we took the Seurat object from the Nagy *et al* dataset generated by the previous section and subset the astrocytes. We then removed the integrated assay and removed two batches as they had fewer than 100 nuclei (1M and 2M) and we were unable to successfully integrate them with the larger batches. We then subset the count matrix to genes within the FADS1_DLPFC_ network and ran the standard Seurat integration and analysis pipeline as previously described. We used the first 10 principal components and a resolution of 0.2 to generate clusters and a UMAP dimensional reduction of the nuclei. We then extracted the metadata with the FADS1 based clusters and combined it with the full astrocyte count matrix to perform downstream analysis, such as identification of cluster marker genes and differential expression.

### Differential abundance testing

We conducted differential abundance testing between control and MDD cells using the miloR v1.2.0 package ^93^. We started with our FADS1 clustered object containing the full count matrix and converted it to a Single Cell Experiment object and subsequently to a Milo object. We used the following parameters for the analysis; k = 20, d = 20 and prop = 0.2. We then tested neighbourhoods for differential abundance using batch and age as covariates.

### Spatial analysis of module scores

We downloaded 10X Visium data from Maynard et al (samples: Br2743, Br2720, Br3942, Br6432, Br6471, Br6522, Br8325, Br8492, Br8667) ^32^. We normalized data using the Seurat pipeline and the SCTransform() function. We created a module score using the AddModuleScore() function. This score was created using all genes enriched in broad cell types or the FADS1-defined astrocyte clusters. Marker genes were those with a positive logFC and an FDR P-value < 0.05. Correlations between cell types were generated independently for all slices. We then averaged the correlation coefficients of all slices to obtain the final correlation matrix for plotting.

### Pseudotime analysis

We conducted pseudotime analysis using the monocle3 v1.3.1 package ^94^. We converted the FADS1 clustered astrocyte object with full count matrix to a cell data set object and used the Seurat UMAP embeddings to learn a pseudotime trajectory using standard settings. Considering there was a notable decrease in Cluster 1 astrocytes in the MDD group and our question was if we could identify a transitional trajectory away from this cluster, we designated Cluster 1 as the root of the graph. We then identified genes differentially expressed across pseudotime using the graph_test() function. We conducted K-means clusters on the top 300 DEGs using the kmeans() function and created a heatmap for expression of these genes using the ComplexHeatmap v2.10.0 package ^95^.

### Cell-cell interaction analysis

We conducted cell-cell interaction analysis using a consensus of methods as implemented in the LIANA v0.1.6 package ^34^. We used the normalized object with all cell types present in the DLFPC for the analysis. We used the liana_test() and liana_aggregate() functions to identify consensus ranks of cell-cell interactions from this Seurat object. We then filtered the results for ligands and receptors that were present in the FADS1_DLPFC_ positive and negative networks, respectively. We also excluded any interactions with an aggregate rank > 0.05 (similar interpretation to a P-value threshold).

### Perturbagen analysis

We conducted perturbagen analysis using the DrugMatrix and Connectivity Map databases using the iLINCS software ^48^. We used the online interface provided and the standard settings to identify patterns of association (**Supplementary Table 8**). Results were returned in the form of absolute Z-score. We aggregated the significant results by perturbagen to identify targets associated with multiple concentrations or cell lines.

### Open Targets

We used the Open Targets Genetics platform ^96^ to search for GWAS hits associated with PPARA. We conducted our search in December 2022. We manually annotated the results into broad groups to facilitate interpretation. We provide the data using the Open Targets L2G score, which is analogous to a probability score based on a machine learning pipeline trained to identify likely causal genes from significant GWAS hits. The L2G score can be interpreted as probability the gene is the target of the causal locus under the assumption the GWAS is similar to those included in the training set.

### Statistical analysis

All analysis unless otherwise stated were carried out using R v4.1.1 ^97^ and Rstudio v1.4.1717 ^98^. GWAS enrichment using MAGMA or H-MAGMA was conducted using the MAGMA command line software. All package versions are specified in their relevant methods section. Plots were generated using either the packages mentioned throughout the methods section or in ggplot2 v3.4.0 ^99^. Plots were combined into figures using Inkscape v1.0. A significance threshold of 0.05 was used throughout, with the Benjamini-Hochberg method (False Discovery Rate; FDR) used to correct for multiple comparisons. The relevant statistical tests used are described throughout the results section and the figure legends.

## Data Availability

All data are available from the studies referenced throughout the text or as source data files.

## Acknowledgements

This work was funded through a Hope for MDD Research Foundation grant to Dr Michael Meaney. The Genotype-Tissue Expression (GTEx) Project was supported by the Common Fund of the Office of the Director of the National Institutes of Health, and by NCI, NHGRI, NHLBI, NIDA, NIMH, and NINDS. The data used for the analyses described in this manuscript were obtained from: the GTEx Portal or dbGaP accession number phs000424.v9.p2. Schematics were created with BioRender.com.

## Author contributions

EF conceived the study, with input from MJM. EF, NOT and IP analysed the data. EF interpreted the data and wrote the manuscript with input from GT, CN and MJM. All authors approved the final version of the manuscript.

## Figure legends

**Supplementary figure 1**

A. Enrichment of MDD TWAS genes in WGCNA modules using a Fischer’s Exact Test. Each point represents an individual WGCNA module tested for enrichment. Dashed line is the Bonferroni threshold for multiple comparisons.

B. Enrichment of MDD TWAS genes in MDD GWAS (Howard *et al* 2019) using MAGMA with 3 different windows and H-MAGMA, using Hi-C data from the DLPFC.

C. Enrichment of genes present in >5 (orange) or <5 (blue) networks within the MDD GWAS from Howard *et al* (without 23 and me subjects) using H-MAGMA. Across all panels the dashed line represents the threshold for statistical significance.

**Supplementary figure 2**

Alternative Mendelian randomization methods (y-axis) for an effect of FADS1_ACC_ or ZKSCAN8_ACC_ network eQTLs on suicidal ideation. The estimate of the effect size with 95% confidence intervals are presented on the x-axis. Dashed line passes through zero.

**Supplementary figure 3**

A. Size of FADS1 and ZKSCAN8 bootstrapped networks across 11 regions of the brain and spinal cord, dashed line indicates the median network size across all regions. *FADS1* median = 823 and *ZKSCAN8* median = 1122.

B. There are no changes in expression of *FADS1* or ZKSCAN8 between males/females or high/low MDD polygenic risk scores (PRS). Panels are labelled with the gene and region for which the relevant expression comparison is conducted. Comparisons were made using a Wilcoxon test.

**Supplementary figure 4**

Protein-protein interactions for members of the FADS1_ACC_, FADS1_DLPFC_ and ZKSCAN8_ACC_ networks from STRING. Each node represents a protein in the network with edges representing protein-protein interactions. Nodes are ranked based on their connectivity. Each network is significantly more connected than expected by chance (FADS1_ACC_ enrichment P-value < 1.0e-16, average node degree 12.4; FADS1_DLPFC_ enrichment P-value < 1.0e-16, average node degree 10.4; ZKSCAN8_ACC_ enrichment P-value < 1.0e-16, average node degree 11.8)

**Supplementary figure 5**

A. UMAP coloured by batches.

B. Stacked bar plot showing the proportion of nuclei in each cluster by batch.

C. Astrocytes clustered using standard methods but coloured by their cluster designation from the FADS1_DLPFC_ network clustering.

D. Differentially abundant neighbourhoods stratified by the astrocyte cluster in which they predominately reside. A negative Log fold change (and red colour) indicates a decrease in abundance in MDD.

E. Enrichment of cluster marker genes in a layer 1 astrocyte signature derived from Batiuk *et al*’s ^33^ mouse dataset and astrocytic genes, whose protein product is present at the tripartite synaptic structure (as identified in mice by Takano *et al* ^100^). Larger dot indicates P-value < 0.05, smaller dot indicates P-value > 0.05. A darker brown colour indicates a smaller P-value.

F. H-MAGMA enrichment of markers of astrocyte clusters in a MDD GWAS.

Enrichment of cluster marker genes in a layer 1 astrocyte signature derived from Batiuk *et al*’s ^33^ mouse dataset and astrocytic genes, whose protein product is present at the tripartite synaptic structure (as identified in mice by Takano *et al* ^100^). Larger dot indicates P-value < 0.05, smaller dot indicates P-value > 0.05. A darker brown colour indicates a smaller P-value.

G. GO terms enriched in cluster identifying genes of Cluster 0, Cluster 1 and Cluster 3.

**Supplementary figure 6**

A. Overlap between FADS1_ACC_, FADS1_DLPFC_ and ZKSCAN8_ACC_ negative networks.

B. Clustering of excitatory neurons by FADS1_DLPFC_ negative network identified 10 clusters.

C. UMAP coloured by Control and MDD.

D. UMAP coloured by batches.

E. FADS1_DLPFC_ negative network module score across all FADS1_DLPFC_ negative network clustered excitatory neurons shows high levels of expression in all clusters.

**Supplementary figure 7**

A-C. PPARA is co-expressed with FADS1 in the DLPFC and ACC, and ZKSCAN8 in the ACC (A). Neither PPARD (B) or PPARAG (C) is co-expressed with FADS1 in the DLPFC and ACC, and ZKSCAN8 in the ACC.

