## Supplementary figures and images for "Mechanistic convergence of depression and suicidality on astrocyte fatty acid metabolism"

### Figure S1

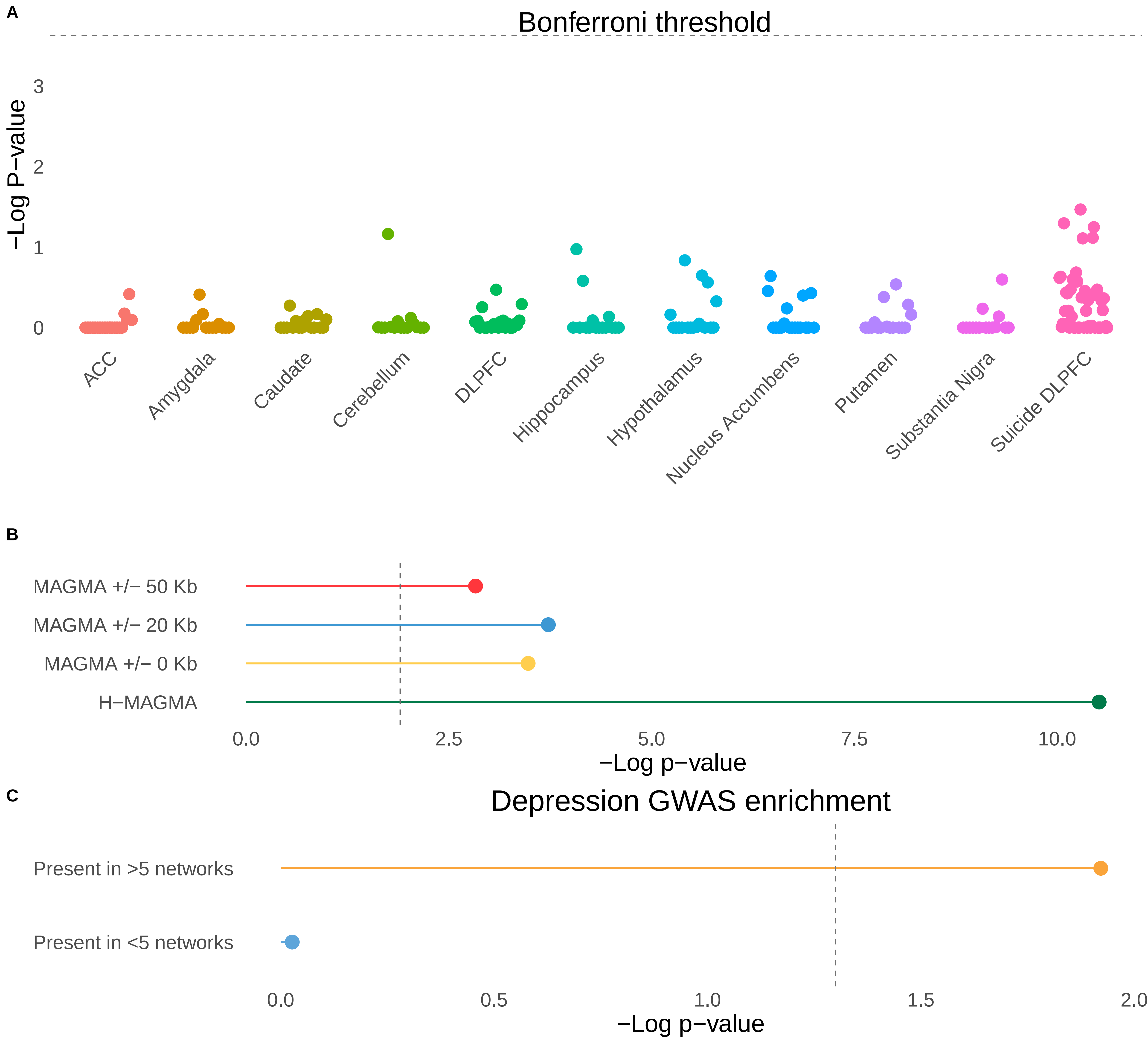

### Figure S2

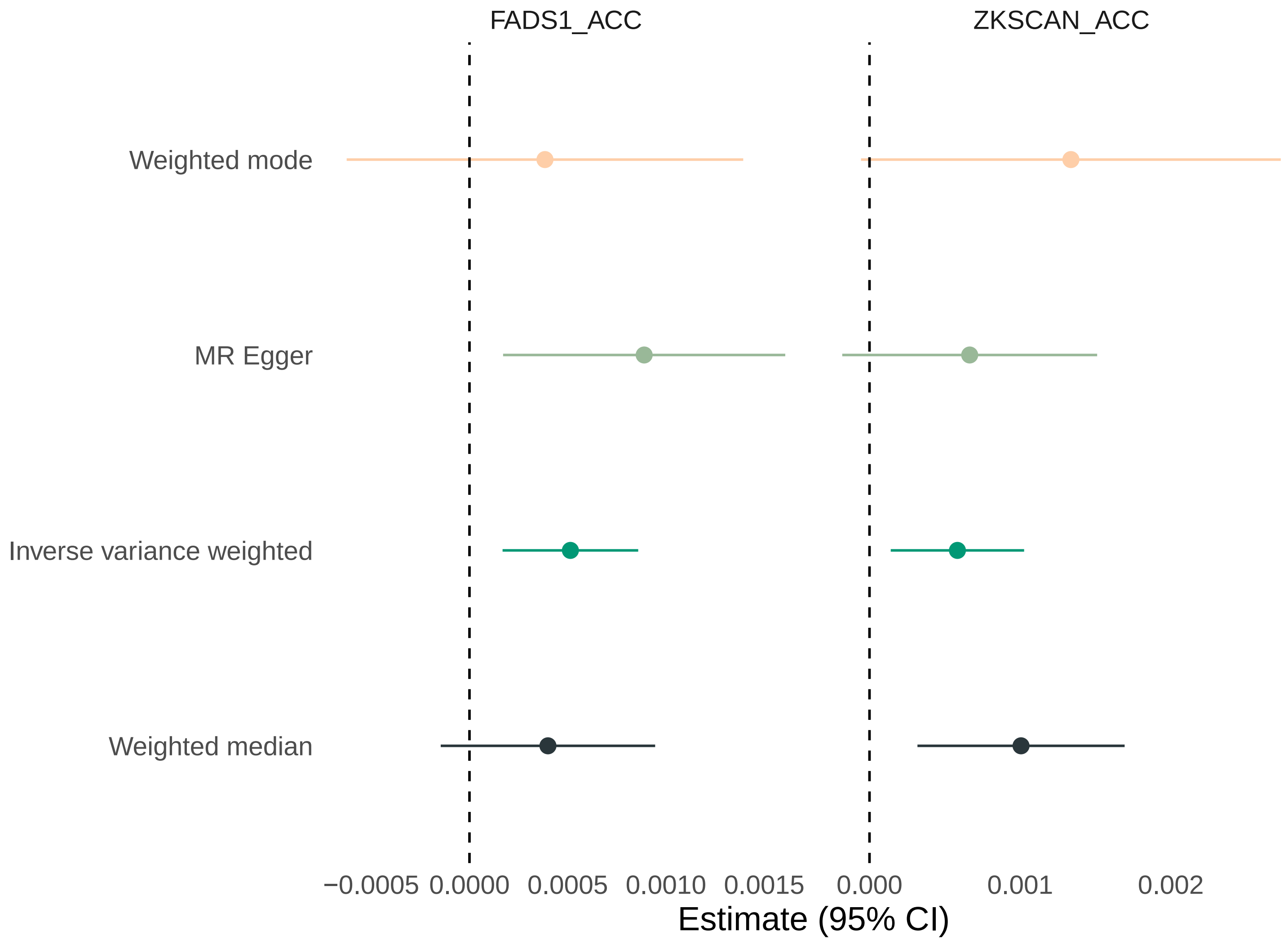

### Figure S3

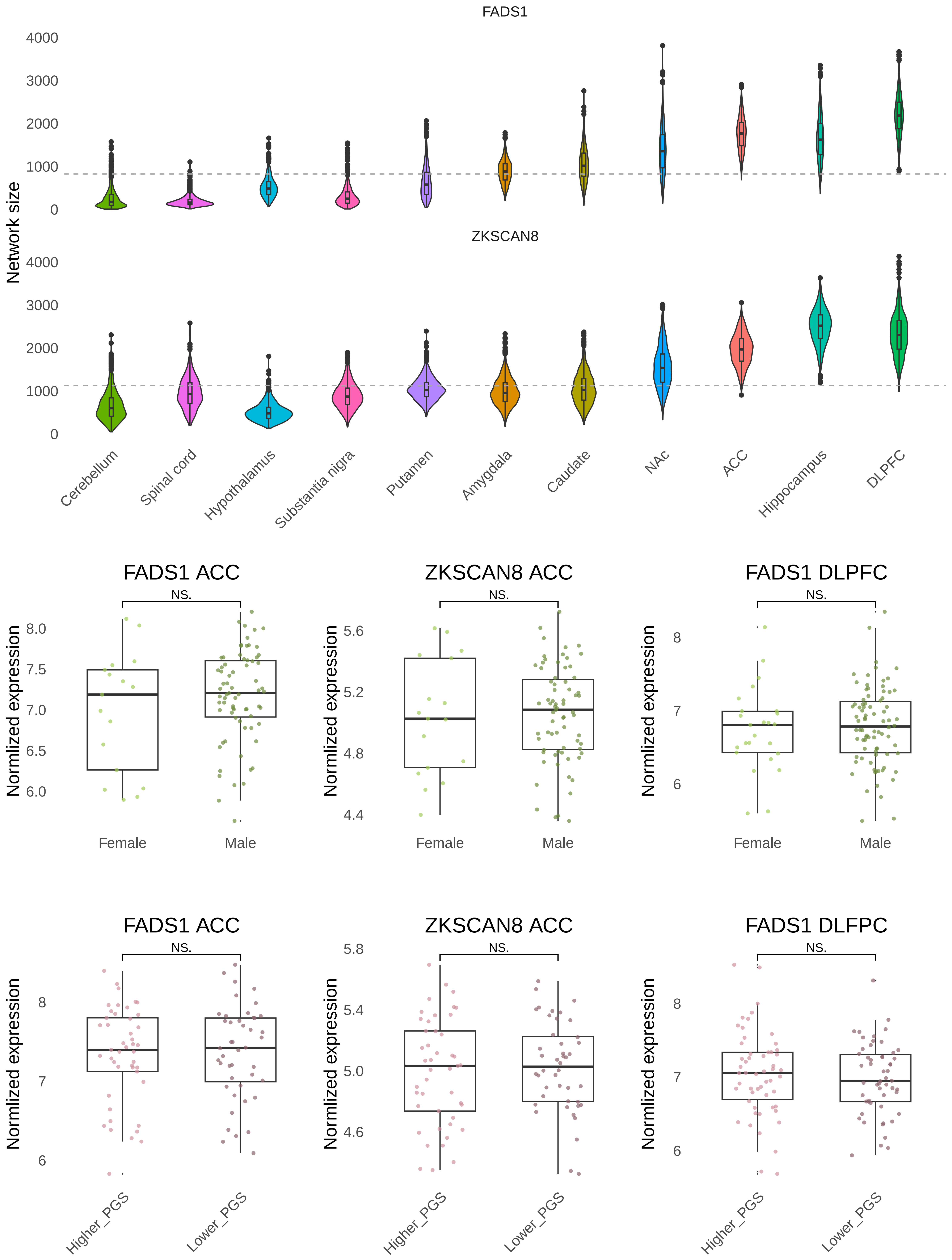

### Figure S4

#### Supp figure 4

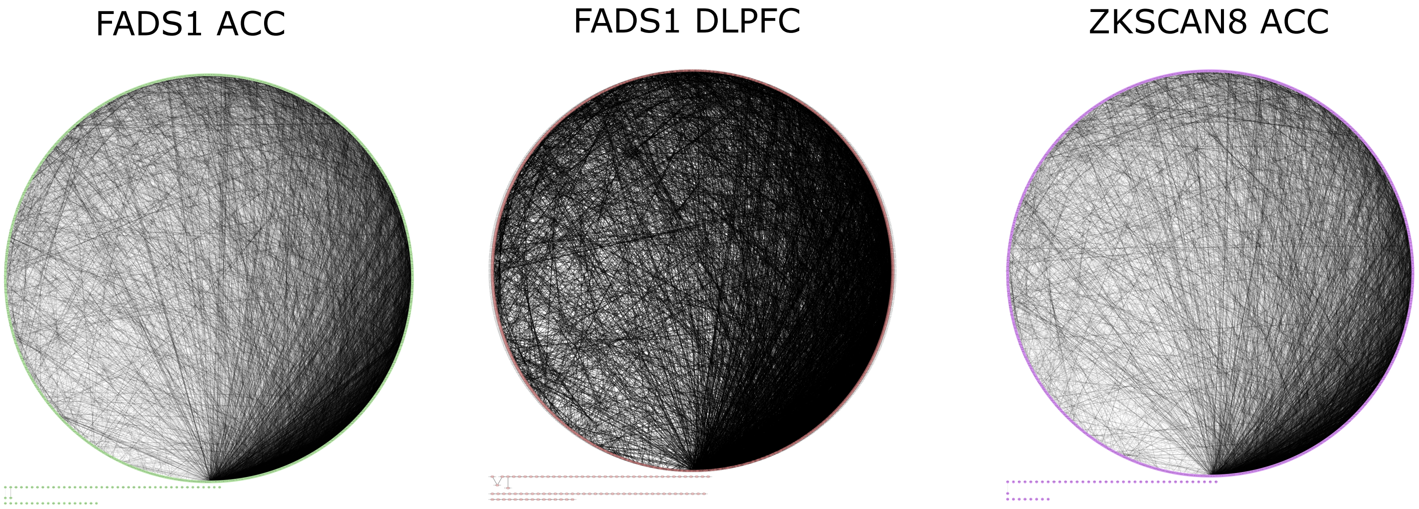

### Figure S5

#### Supp figure 5

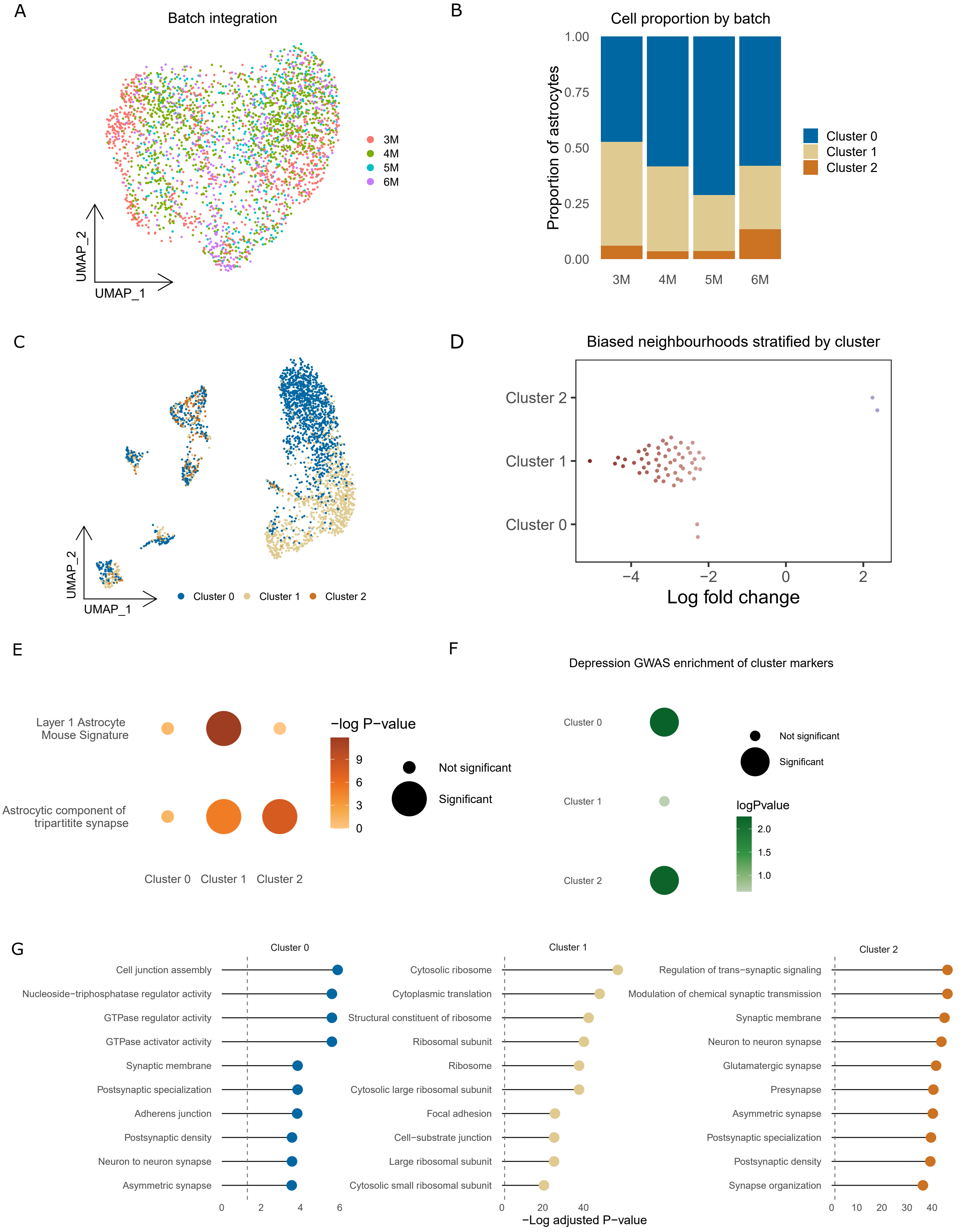

### Figure S6

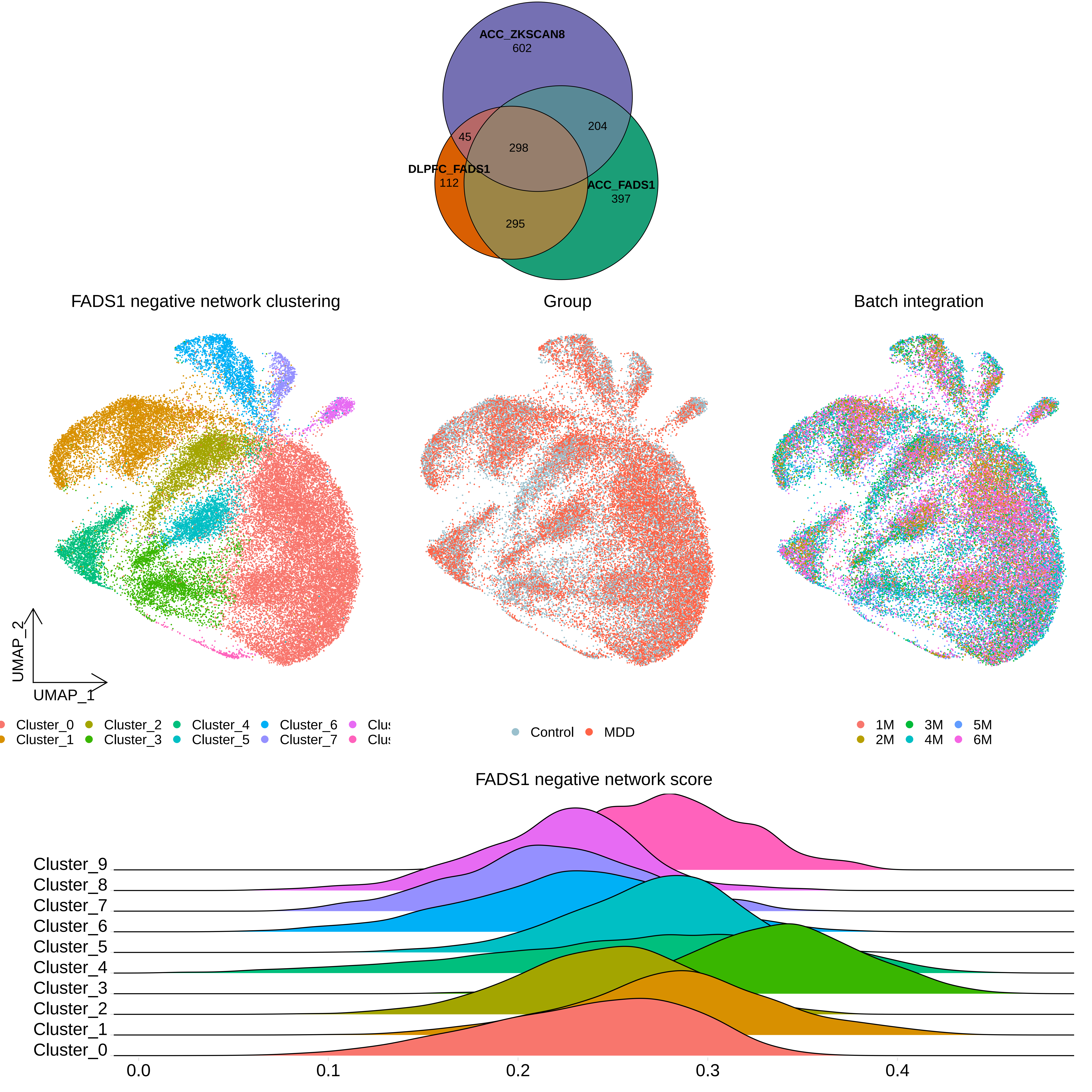

### Figure S7

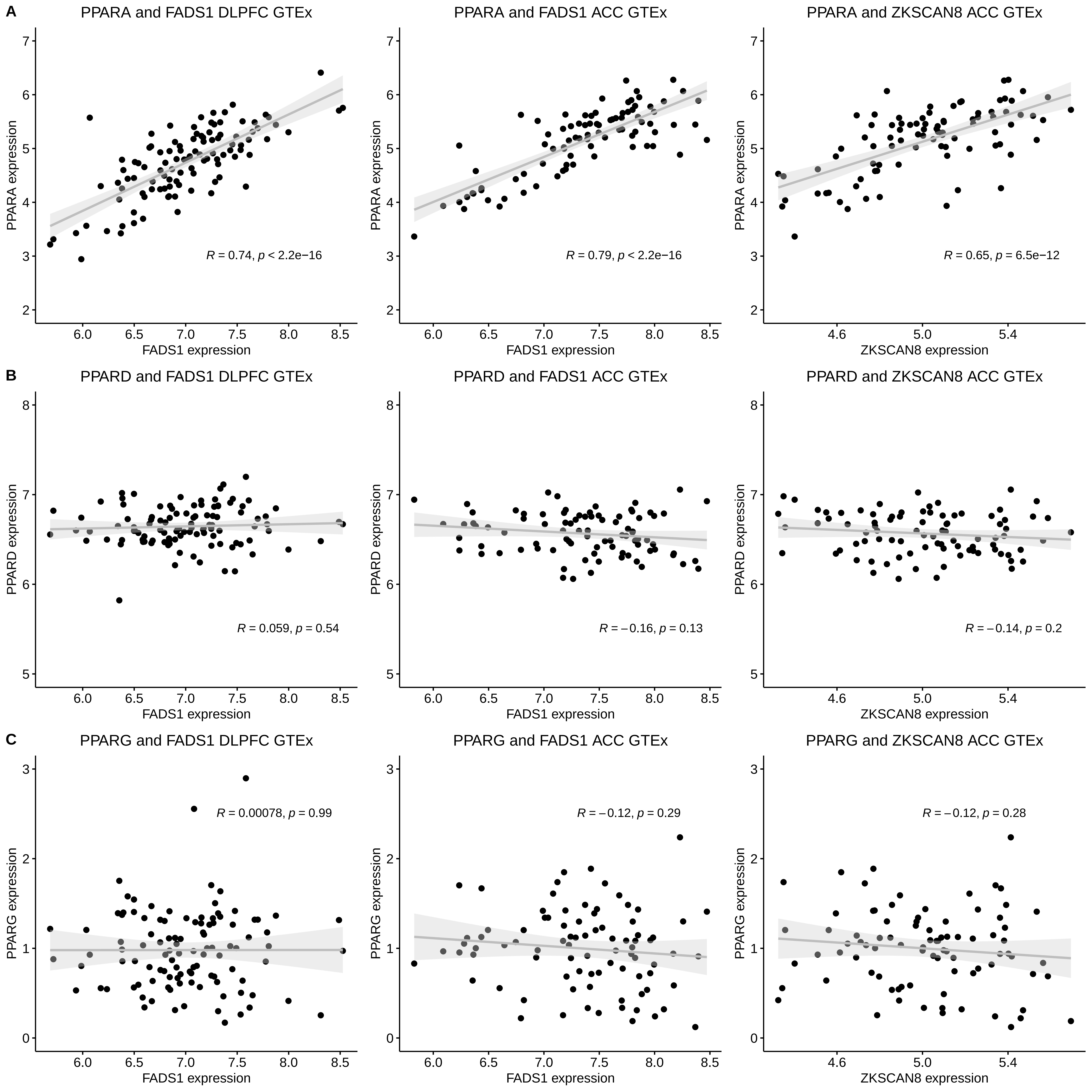
